# Health financing in Sudan: key informant interviews in the wake of the 2023 conflict

**DOI:** 10.1101/2023.12.20.23300333

**Authors:** Fatima Bashir, Luke N Allen

## Abstract

**Background:** Sudan is a large, landlocked, African country with a population of 46 million. It is considered a ‘least developed’ country, and coverage of essential health services is low. In April 2023 fighting broke out between the Sudanese Armed Forces and the paramilitary Rapid Support Forces. The conflict quickly escalated and has claimed 9,000 lives so far, with hundreds of thousands of people internally displaced. It is unclear exactly how the war has impacted Sudan’s already fragile health financing system.

**Methods:** We conducted one-to-one semi-structured interviews with a purposive sample of experienced health financing policymakers and policy advisors working in- or with the Sudanese government. Five senior key informants were recruited using snowball sampling. Interview transcripts were analysed using thematic analysis, with reference to the WHO national health financing framework and Sparkes and colleagues’ political economy framework.

**Findings:** Conflict has undoubtedly undermined Sudan’s ability to mobilise, allocate, and distribute financial resources. However, the existing health infrastructure was already so weak, and state funds so meagre, that the challenges posed by fighting were overshadowed by themes around Sudan’s underlying structural issues: out of pocket payments account for over 70% of all health expenditure; the existing national insurance scheme only covers a minority of the population; and there is unclear delineation of responsibilities between purchasers and providers and between the federal and state ministries of health. Major themes emerged around governance, revenue raising, risk pooling, purchasing and service delivery, and external donor funding.

**Conclusions:** Investment in developing health financing capacity at the state level, implementation planning, blended financing for the national health insurance fund, donor coordination, and community mobilisation to agitate for greater risk pooling were identified as essential steps. Without robust mechanisms to pool risk and diversification of financing sources, it will not be possible to extend financial risk protection or basic health services to large sections of Sudan’s population.

## Introduction

Fighting broke out across Sudan in April 2023, severely impacting an already fragile health system that serves one of the worlds’ least developed countries. Even before the war, Sudan’s life expectancy at birth was only 65.3 years (1) and its under 5 mortality rate was double the global average (54.9 deaths per 1,000 live births).(2) The estimated number of people who need humanitarian assistance reached 24.7 million in May 2023, representing almost half of Sudan’s total population. WHO estimate that 11 million of these people require emergency assistance for life-threatening conditions or life-sustaining support to meet minimum living standards (3). These figures will continue to rise as the ongoing conflict progresses.

Anecdotal reports suggest that Sudan’s health financing system has been crippled by the conflict, severely impeding service delivery at a time when it is needed most. However, it is not clear which parts of the system are under greatest stress, or what actions should be prioritised to safeguard financing of essential health services. There is an urgent need for information around exactly how the conflict has affected Sudan’s health financing situation and what actions should be taken in the short and longer term to alleviate critical pressures - and build a fairer and more sustainable system for the future.

In this paper we aimed to evaluate Sudan’s current health financing situation, exploring the perceptions of health finance experts on how the conflict has impacted the current model, and identify priority actions to secure health financing for essential health services. We present an extended background review of the context and current financing landscape before presenting methods for key stakeholder interviews, findings, and recommendations.

### Geopolitical context

Sudan is the third largest country in the African continent and has an estimated population of over 46 million. According to UNCTAD it is considered a ‘least-developed’ country (4) and continues to be afflicted by protracted conflict, regional inequities, and is considered a fragile state struggling to perform basic essential functions to meet the basic needs of its population. The economic situation in Sudan had seen a slow but gradual increase in GDP by an average of 6% per year from 1999-2010, until the succession of South Sudan in 2011. The World Bank Human Capital Index ranks Sudan 139 out of 157 countries. Approximately two thirds of Sudanese are living below the national poverty line, with the highest poverty rates observed in conflict-afflicted states (5). A third of the population do not have access to safe drinking water sources (UNICEF Sudan Water, Sanitation and Hygiene Annual Report, 2020) (5, 6). Just over 60% of the population are literate, and the average citizen has 3.8 years of schooling. Life expectancy is 65.3 years and the leading causes of death are pneumonia (7.4%); followed by malaria (6.8%); and then malignant neoplasms (4.6%) (5).

Sudan has long experienced political instability. The most recent coup occurred on October 25, 2021, when the Sudanese armed forces (SAF) and the paramilitary Rapid Support Forces (RSF) deposed the transitional government (7). Strain between the RSF and SAF had been growing alongside political negotiations on the merging of both forces. Tensions escalated into a full-scale armed conflict on April 15, 2023, when both warring factions exchanged intense fire north of Khartoum which quickly spread across the country (8).

Sudan’s poor relations with the international community has made it ineligible for debt relief. Access to grants and funding from global and regional financial institutions for development initiatives has also significantly impacted the population’s health (MICS, 2014) (6). There was considerable effort being made on this front under the leadership of the transitional governments’ Prime Minister Hamdok, however these efforts came to an abrupt end with the October 2021 coup (9).

### Sudan’s health system

In theory, the health system in Sudan is decentralized and mirrors the overall political system with three levels of governance: federal, state, and locality. However there is an absence of a clear demarcation of the roles and responsibilities of each tier (10, 11). The Federal Ministry of Health (FMOH) is home to ten general directorates, and is mandated with health sector financing, policy-making, strategic planning, technical support, and international/global relations, (10). Eighteen state ministries of health (SMOH) are responsible for the implementation of policies and service delivery at district level.

Policy is usually formulated by the FMOH However, the WHO Joint Annual Review of 2017 noted that some states do not use these national policies and guidelines - particularly for primary care units. This review also highlighted the inequality of access and uptake of services among and within the states. Sudan’s universal health coverage (UHC) service coverage index was reported at just 44.3% in 2017 (5). While government reports emphasize high accessibility to primary care facilities (specifically within 5 km reach), most facilities are unable to provide all essential health services (11, 12). The health system is considerably hospital-centric, with much fragmentation and inefficiency due to the proliferation of providers and stakeholders in the absence of coordination or clear planning.(5)

Sudan’s human resources for health (HRH) is facing several challenges. There is considerable disparity in the distribution of HRH between public and private sectors and between urban and rural areas (5) Additionally, ‘brain-drain’ (the migration of trained medical professionals outside Sudan) and high turnover of staff remain constant threats to the capacity of the FMOH. According to the World Bank’s latest estimate, there are 1.1 nurses and midwives per 1000 population, and 0.3 physicians per 1000 population (1), in comparison to the WHO recommendations of 4.45 physicians, nurses, and midwives per 1000 population, these figures are inadequate for achieving UHC (13). Moreover, 70% of the health workforce currently work in urban settings despite 70% of the population residing in urban areas (14).

### Overview of health financing landscape

The health sector suffers from limited financial resources. The main financing sources are the Ministries of Health, the National Health Insurance Fund (NHIF), the Armed Forces Health Insurance Schemes, out of pocket (OOP) spending, and international aid/donors’ funds. Health Expenditure was reported at USD 2.4 billion in 2018 (USD 58.84/capita); down from USD 4.8 billion (USD 132.3 per capita) in 2015.

Health facilities at different levels receive funds from different sources, each with different and distinct incentive structures. External aid funds are managed by the FMOH and are used to finance vertical programs such as HIV, TB, and malaria. Staff salaries are managed by the SMOH, while curative care is funded by the NHIF, the Ministries of Health, and the Ministry of Welfare and Social Security (12). Delivery of health care services is relegated to the 18 states, while facing formidable challenges in mobilizing adequate financing for their functioning. A significant share of the federal consolidated health budget is transferred through federal block grants with lack of clarity of laws that delegate responsibility and financing functions to states and localities (12).

Public expenditure on the health sector has remained at between 7-8% as a share of total public spending during the last decade, representing about 1.5% of GDP (11). Despite the Government commitment to providing free emergency medical care, free medicines for children under 5, and free medical care for specific diseases, a considerable amount of OOP expenditure is still incurred: 74% of total expenditure on health in 2018, reflecting a small decrease since 2015 (83.3%), but still far above 2008 levels (63%).(5) While OOP represents the main component of health expenditure, the role of donor funds is catalytic in certain areas, particularly those related to strengthening the health system.(15) Development partners including GAVI and the Global Fund for AIDs, TB, and Malaria (GF) support multiple health systems ‘building blocks’, including HRH capacity building, procurement, and supply chain management system, and health information systems in Sudan (15). The National Health Sector Partners’ Forum is the main coordination mechanism for all the health and development sector partners.

Figure 1 shows that overall health spending was rose between 2002 and 2008, mainly driven by an increase in government, pre-paid, and DAH spending. Government spending fell after 2010 but overall health expenditure remained at around USD 2 billion, with the shortfall covered by an expansion in OOP and a small absolute increase in DAH.

**Figure 1:**
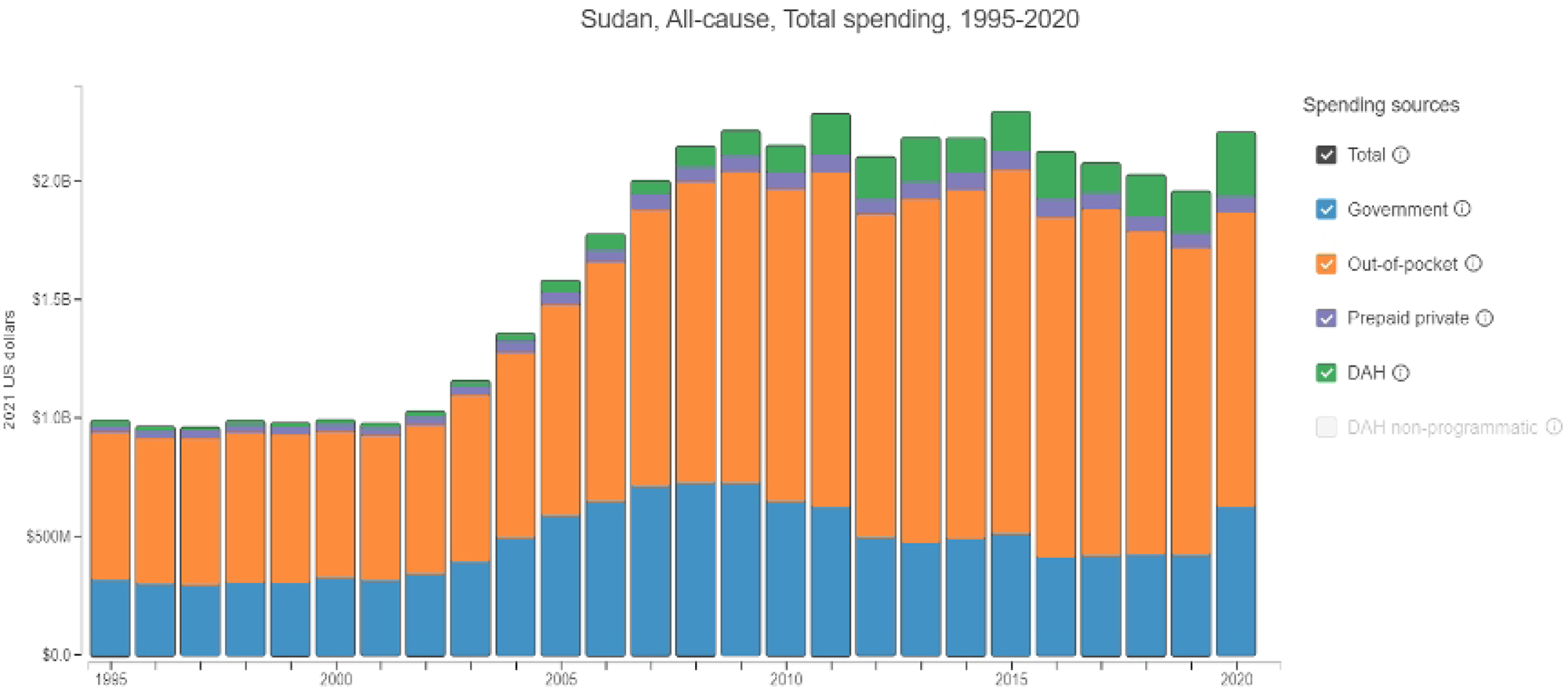
Trends in total health expenditure *Source: Institute for Health Metrics and Evaluation vizhub*

Major financial protection schemes include the NHIF which covers approximately 43.8% of the population (nhif.gov.sd, 2016), and premiums for households living below the national poverty line are covered by public funds (12). While there are admirable attempts to expand coverage, the health financing landscape is impeded by fragmentation of funding pools, especially those of the FMOH and NHIF and the overall lack of coordination and management capacity (12). There are other religious-based social security taxes, such as the Zakat Fund, which provide further support and subsidize the health care costs of lower income groups, however these only cover small minorities of the total population.(12) In terms of the main health financing challenges facing Sudan before the current conflict, the overreliance on OOP created inequities in terms of access to treatment and care for vulnerable populations.

### The October 2021 Coup

On October 25, 2021, the Sudanese General Commander Abdel Fattah al-Burhan announced the dissolvement of the transitional government and Sovereignty Council, declared a state of emergency, and placed Prime Minister Abdalla Hamdok and most of his cabinet under arrest (16). While the military power termed this endeavour as a ‘correction of the political pathway’ it was in effect a coup d’etat and was recognized as such by most of the international community.

The coup affected the health system in many ways; Alhadi et al summarised these impacts in the following: creation of a leadership void: both Federal Minister and undersecretary were dismissed in the immediate aftermath of the coup effectively terminating leadership and lines of command which led to a severely felt disorientation in terms of management and coordination; interruption in donor funding: on a background of difficult major financial reforms that were taking place in the country prior to the coup, post-coup events further exacerbated the financial situation with major donor funding being frozen in a direct response to the military takeover. The USA alone suspended over USD 700 million of its direct assistance to Sudan’s government (7, 17). The suspension of Sudan from the African Union (18) created further challenges around the country’s ability to access external aid through regional development funds (access is often contingent on AU membership). This suspension also led to the terminating of funding from the World Bank. This withdrawal of funds severely impacted the health sector - particularly in the capital Khartoum where 70% of health services are delivered. Furthermore, medical professionals have been a political target for security forces as they are considered significant challengers for military rule (7). There is a lack of clarity on how the coup directly affected financing structures and what mechanisms were put in place to absorb the suspension of external aid. There have not been many economic evaluation studies conducted examining the economic effect of the coup on OOPs or other financing schemes.

### The April 2023 War

The violence that has erupted in Sudan on April 15^th^ was triggered by a power struggle between the leaders of the SAF and RSF. The impacts on the health system have been dire and may constitute war crimes. The UN has estimated that 9,000 lives have been lost (19, 20). As of October 18th, 2023, WHO has verified 58 attacks on health care of which 31 sustained deaths and 38 injuries of health workers and patients have been verified by WHO (21). The Sudanese government’s ‘Combatting Violence Against Women Unit’ has reported that 88 incidences of conflict-related sexual violence have been verified(22). We feel these figures are very likely to be underestimates. The WHO has stated that 70% of health facilities located in areas affected by fighting are out of service leaving only one third of hospitals operational in conflict zones.(23) In Khartoum, the most affected region, only 20% of hospitals fully functioning and 43% are partially functional (24, 25).

The situation is further complicated by power shortages, looting and vandalism of facilities, limited medical supplies, and damage to critical infrastructure hampering the delivery of essential health care. The disruption of the national vaccination programme comes amidst an ongoing polio outbreak. The majority of cold chain facilities have either been looted, damaged and/or destroyed, leading to the loss of over 600,000 vaccines. The nutrition crisis is dire; UNICEF has reported that Sudan currently has one of the highest rates of child malnutrition anywhere in the world (26).

This recent conflict has resulted in an estimated loss of USD 700million to its already underfunded health system (23). As the was continues to ravage the country, the full effects on the health system remain to be fully seen. This review revealed several gaps in the information and data available on the financing structure and its response to the crises. We attempt to fill these gaps with the findings from the interviews of key informants and experts.

## Methods

### Reflexivity

FB is a female Sudanese clinician and global health programme and policy professional who was undertaking her Master’s in Health Policy at Oxford’s Blavatnik School of Government during the project. She has experience working within the Sudanese health system as well as working at a number of UN agencies. LA is a male British clinician who works as a global health researcher and policy adviser in a wide range of high-, middle-, and low-income settings. He has led numerous national mixed-methods health system assessments for WHO and the World Bank, and has previously worked with health financing policymakers in Sudan. Both authors have masters-level training in qualitative research methods and previous experience conducting interviews.

### Sampling and recruitment

We obtained a purposive sample of leading health financing policymakers and policy experts who have first-hand experience of working with or advising the national Sudanese Federal Ministry of Health (FMoH) before, during, and after the onset of the April 2023 conflict. Whilst up to twenty people meet these criteria in terms of their job title, we excluded those with no formal training or expertise in health financing. We used a snowball approach to recruitment, starting with two senior national health financing policymakers and an internaitonal health economic advisor who has supported the FMoH; all of whom were already known to us in our professional. We informed all potential interviewees about our aims and rationale, as well as our personal interest in the subject which relates to securing sustainable health financing in order to equitably extend access to essential health services via email. We aimed to perform sampling and analysis iteratively, stopping recruitment when we achieved thematic saturation (27, 28). As it was, our key informants only recommended two additional interviewees and no further suitable informants were found or recommended, relecting a critical dearth of expertise in Sudan. As such, our sample size contained five key informants; a mix of men and women. No invited key informants refused to participate.

### Modality

The 45-60 minute interviews were conducted remotely via the Zoom video-calling platform. Interview recordings were translated, when necssary, from Arabic to English and transcribed. Both researchers conducted the first three interviews jointly, and FB conducted the final three on her own. Noone else was present in the interviews except the participants and researchers. Field notes were taken during the interviews. All interviews were primarily conducted primarily in English; the professional working language for all interviewees. We did not conduct repeat interviews and transcripts were not returned to participants for comments or corrections. Participants did not check the findings.

### Analytic framework

We adopted a phenomenological approach as we were interested in understanding the experiences and perceptions of our key informants. We used the WHO national health financing framework to structure our analysis. The framework was developed in 2017 by WHO with the objective of providing support on health financing policy to its Member States to develop national health financing strategies, highlighting the different aspects of health financing policy which need to be analysed and addressed by countries from a universal health coverage (UHC) perspective. The WHO framework highlights the connection between health financing and overall health system objectives, directly and indirectly via the intermediate objectives (in Fig. 1). Of relevance to this study, the framework emphasises that the health financing system does not act alone in affecting the intermediate objectives and final goals; and rather requires coordinated policy and implementation strategies across the various health system functions.

The framework combines a normative set of goals that are embedded in the concept of UHC with a descriptive framework of the functions and policies that are part of all health financing arrangements. For the purpose of this study, our normative position is analysing the current financing structure and its response to crisis while exploring policy options that drive future reforms in health financing in line with achieving UHC. As such, our analysis was guided by the main four components on the left of the depicted framework: revenue sources and contribution mechanisms; pooling of funds; purchasing of services; policies on benefit design; and governance structure. We used these elements analyse the current situation of the Sudanese health system financing landscape and how this is structured between federal and state levels; how the different sources of revenue/pooling mechanisms and benefits structures are adapted/utilised in times of political and natural crisis; and what governance structure guides this process.

**Figure 2:**
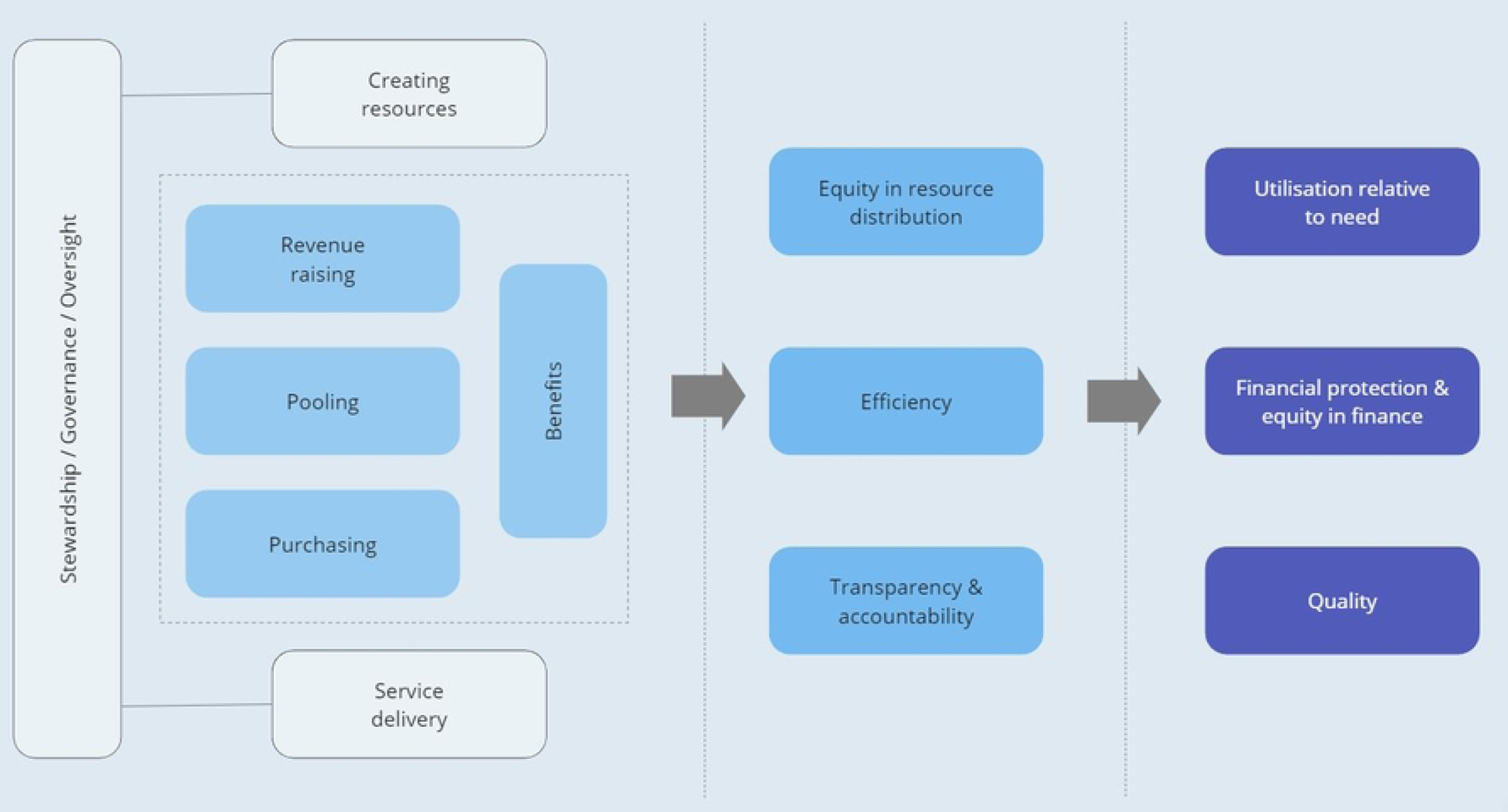
WHO national health financing framework

We also drew from the Sparkes and colleagues Political Economy Framework (PEF) for Health Financing Reform which builds on the methods of applied political analysis and strategies to assess the power and position of key political actors to develop strategies to alter the political feasibility of desired reform outcomes. This framework can be used to analyse health financing reforms based on the roles and positions of different stakeholder groups and political structures of relevance.(29) We used this framework to identify players, their interests and motivations, and their power as we explored our key informants’ perspectives of the effects of political volatility in Sudan on health financing structures – and the feasibility of any suggested policy recommendations.

### Topic guide

Based on the Political Economy and WHO national health financing frameworks, we developed a semi-structured topic-guide around seven open-ended questions. We used probing questions to follow up on cues. The guide was not piloted. Data were collected from June to September 2023.

#### Current structure and impact of events

- What is the current health financing situation; and how does it vary by state?
- What has happened to health financing in Sudan since April 2023?

#### Governance

- What are the governance structures that guide the allocation between states and in times of crisis?
- Who are the major actors? what are their interests what power to they yield?

#### Reform

- From your experience, what would be a sustainable refom policy that could be further explored?
- How feasible is this suggestion?
- What political enablers need to be in place for this to succeed?

### Analysis

We used a blend of inductive and deductive thematic analysis, with reference to the PEF conceptual framework and the WHO financing framework. The flexibility of thematic analysis allowed us to consider both manifest and latent content in data analysis. Analysis was conducted following the six steps outlined by Braun and Clarke; beginning with familiarizing the data, followed by generating initial codes, searching for themes, reviewing themes, defining, and naming themes and finally producing the final report.(30–32). FB led the analysis and LA independently coded three transcripts. Preliminary findings were discussed and refined iteratively. We used NVivo Open Code 4.03 software.

### Ethical considerations

The study was carried out in accordance with the principles of the 1975 Helsinki Declaration. Participation was voluntary and informed written consent was obtained from all participants prior to participating in the interviews. Ethical approval was granted by the LSHTM research ethics committee. No research ethics committees are currently operational in Sudan. Ethical approval was granted on 7^th^ August 2023 at 14:57 and the first recruitment email was sent at 10:17pm that same day. Recruitment ended on the 25^th^ August 2023. Given the small number of informants and the potential risks involved if their identities were to be revealed, we do not present their specific job roles or personal characteristics, neither so we attribute quotes.

## Findings

### 1. Governance for Health Financing

#### 1.1 Implementation challenges

Whilst health financing reform has been high on the agenda of most stakeholders implementation has remained a key challenge. Sudan has financing plans and policies, as well as well-trained staff and administrators in many regions and levels of the health system. However, indistinct lines of accountability and that lack of consensus around what each team is responsible for stymies operationalisation.

> *“In 2016 we developed and disseminated the National Health financing policy, which is stated clearly, the functions and the role of each level. But I think like other policies and strategies, we have the implementation challenges regarding the this clarity of all our responsibilities”*.
>
> *“We have many forums. We have many joint committees. We have technical working groups. But .. we need to add … the word <u>effective</u> for each. Effective communication, effective planning, effective coordination, effective implementation. Because most of our resources are lost at during this inefficiency … all the time you have meetings. But what then then the implementation?”*.

For programs funded by the Government, most programs are centralized and, depending on state capacity, funds are redirected to each respective state. There is no clear mechanism by which this distribution happens and is mostly based on ad hoc requests made by the state.

> *“What I hadn’t appreciated before we went was the lack of consensus around what should be done at state level and what should be done at federal level. How any federal level resources should be shared around to states in terms of a fair shares allocation if you like. How resources should be pulled and who should manage those resources”*.

#### 1.2 Federal vs state Financing Allocation

According to key informants, funding to states does not go through the FMOH but through the Federal Ministry of Finance which then distributes resources to state Ministries of Finance. There is no clear mechanism by which this distribution happens and is mostly based on ad hoc requests made by the state. In states where there is insufficient health system capacity, these requests are less forthcoming, meaning that the provision of funds is inversely proportional to state health system capacity. Capacity at state level is complicated by lack human resources for health (who primarily reside in urban areas and/or are lost to ‘brain drain’) and more importantly, deficiency in managerial capacity.

The resulting financing structure remains opaque with an array of funding sources further complicating the multi-tier system and lacked a clear system and/or policy that demarcates and coordinates the funding lines for each. This creates a diverse picture ranging from very well-developed financing models and revenue raised to poorly funded state systems.

> *“It depends in in a large part It depends on the on the strength of those who are responsible for health at the state level. You know, they, because it’s like a cake or a steak; each one wants to take the biggest part at the state level. So some of our DGs [Director Generals] at the state are really very strong and convincing… Some of the DGs or ministers of health are a little bit weak, some of them shy, some of the ministers of finance are completely against health. It’s not their priority.”*

One participant commented on how this unclear financing allocation mechanism has resulted in a vast disparity between the states.

> *“You’ve almost got all these little different economies that together comprise Sudan. So some were very poor, some states very poor, very poorly resourced, not very strong governance, lots of - even diversity within those states in terms of their capability of raising tax or collecting money, getting payments for any health services. And other states quite mature. In terms of how they were, how they were progressing”*.

### 2. Revenue raising through the National Health Insurance Fund (NHIF)

The NHIF is situated under the Ministry of Social Affairs and collaborates with the FMOH. NHIF revenues come primarily from (compulsory) civil servant tax contributions with a small proportion of contributions from the Ministry of Finance covering ‘poor populations’, and voluntary contributions from those working in the informal sector. There is no clear definition of ‘poor’, leading to a mix of under- and over-coverage depending on the region. Only 55.8% of the population is covered by insurance, mostly from the formal sector and residing in mostly urban areas. This makes NHIF look more like traditional private medical insurance than true social health insurance.

> “*You can’t talk about equity at any how because you are just focusing on people who are working in urban area where the health services are provided. So it looks just like a medical insurance rather than social health insurance…If we cover the informal sector at that time, this means that we are talking about equity because most of this informal sector are distributed in the rural area and they are facing the challenges. But the problem is that the definition of informal sector in Sudan is not clear at all.”*

NHIF funds were originally topped up by the Ministry of Finance (using general tax revenues), but these transfers have stopped since the war started in April 2023, fully exposing the insurance fund to a decline in voluntary contributions. Participants highlighted the lack of incentive for enrolment for individuals in the informal sector where service provision is sub-optimal. Covid-19 and the war have reduced the resources available for improving the service offer in rural areas, which has made it even harder to convince people that it is worth paying for insurance. This vicious spiral of diminishing services leading to diminishing voluntary contributions threatens to completely undermine the NHIF.

> *“So when [the national] Minister of Finance faces this problem and or this challenge is of course when they start to cut the budget due to the armed conflict and even the translation In Sudan, NHIF have been affected immediately. Why? Because they lost more than 70% of their total revenues. It’s a problem.”*
>
> *“After the war, there is not any cash transfer from the Minister of Finance and now in NHIF incur this debt and this loss in most of the States and many providers and many facilities now start to stop the contract with NHIF”*.

Two of the interviewees felt that the future sustainability of the NIHF depended on being able to draw on a blend of revenue streams, not just contributions from public servants.

> *“The best way is the mixed way. Between tax and between insurance, because you see, you can’t rely on the tax base for sure. And even due to these challenges regarding the informal sector, you can’t rely on the insurance. You can’t collect money from all the population. So I think the best way is to have this mix but with, with precaution we have to revisit the inclusion criteria of targeting of this vulnerable group.”*

### 3. Risk pooling

#### 3.1 High OOP and lack of risk pooling

The war has crippled the economy, slashed tax receipts, and led to government spending reallocations away from the health sector. However, our interviewees felt that the greatest threat to Sudan’s medium- and long-term health financing is the lack of risk pooling. Clearly the war has made it much harder to institute pooling reforms, however OOP spending accounted for 74% of health expenditure before the war.

> *“The biggest challenge that I think they was faced then and still faced and will be still faced in ten years’ time is this the risk-shift. So regardless of the conflict. Even if the conflict were resolved now.”*
>
> *“At the moment, the financial protection risk sits with the patient. You know, the level of self-pay is enormous… Through risk pooling and risk sharing and financial protection arrangements. It’s being paid for by individuals and out of their pocket”*.

Participants agreed that change required a significant cultural shift, and that devolution may be an important ingredient in reform.

> *“Shifting so that the government taking responsibility or a national health insurance fund taking responsibility for risk pooling is a huge, huge cultural shift. And that to me is the largest single challenge. That I think will be faced in health financing policy in Sudan*.
>
> *“In the long term I could see a situation where each state had its own, that would be like, you know, there would be some federal funding equalization; funding distribution down to the state level. From central taxation or federal taxation. But you would have much more local ability to build up risk pooling not just across the population but over time which is what you’re talking about. You know, it’s not just about protecting individuals who fall ill. By everybody chucking in, like an insurance model does, you’re also temporarily protecting risk because you’re allowed to save and build up reserves and funds. And I think that could be done at a state level. Without question.”*.

#### 3.2 Equity and the lack of adequate coverage of the population

was an aspect widely covered by the participants. While the NHIF provides coverage for those in formal areas of work (i.e., civil servants), a significant proportion of the population works in the informal sector. The lack of incentives of the NHIF service delivery is a significant barrier to attracting individuals from this group, creating considerable inequity in access to health services.

> *“You can’t talk about equity at any how, because you are just focusing on people who are working in urban areas where the health services are provided.”*
>
> *“We are talking about the primary health care services. It should be, I would like to say, free of charge for everyone in Sudan. This is not the case…unfortunately, and should be completely paid by some, somebody. This somebody is health insurance in Sudan, of course. This is written in our policies, in all our strategic plans, in their, the health insurance fund strategic plan itself and so on”*.

The Sudanese government identified ‘poor’ families as eligible for funding under NHIF and funding for this amendment of the NHIF was provided directly from the Ministry of Finance. However, a recurrently mentioned obstacle to this addition was the absence of a clear definition of what ‘poor’ constitutes. There are no clear thresholds for income levels and/or other criteria to fulfil the ‘poor’ requirement. Considering that a large percent of the Sudanese populations is under the poverty line, that would infer that all Sudanese are eligible for coverage under this eligibility criteria.

> *“So I don’t want to say that health insurance fail but I think that it was very challenging to cover the informal sector after that in 2017 the the the contribution of informal sector in NHIF is very low, very minimum, very minimum, OK. Then the government try to change the policy and start to the target, the poor families. OK, and since 2017. It was very huge influx of poor families or enrolment under the umbrella of NHIF maybe 1 to 2 million household per year greater coverage of NHIF from forty-something-percent up to more than 85%, and this is a current NHIF covered. What do you know at that time we face what many or countries face when you are targeting support families. The inclusion criteria again, what do you mean by poor families? So by the end of the day, all the Sudanese, if they are rich or if they are poor, they are urban or rural. They are involved in free scheme”*.

#### 3.3 Community engagement

Increasing insurance coverage and community participation in the NHIF was a prominent theme among participants who highlighted the need to develop a community-targeted strategy to encourage and attract enrolment of those working in informal sectors and to improve risk pooling. However, this requires a large degree of community engagement and trust building to incentivize enrolment by building momentum from existing grass root and community movements that have been central and instrumental in the covid-19 response and during the ongoing conflict in providing health services and awareness raising.

> *“for me the biggest challenge is the lack of risk pooling that to me is fundamental issue. I mean it’s such a huge, huge It’s such a huge disconnect… That even if you were to sort of push for a social insurance type fund or a push for the National Health Insurance Fund to be, you know, all-encompassing with a safety net provided by government funding. You know, all the things we recommended. You’d have to have Such a strong community”*.
>
> *“During NHIF we try to use the communities to motivate them to encourage them to enrol in the NHIF basically, and sometimes we try to give them tasks like a we can recruit the doctors, but we don’t have residence for him, for example. So as community we can give him house or anything like that and and usually usually especially in the rural area we we we realise a very strong commitment and response from the communities because now … they are able to calculate what they will gain and all they lost. So the community is a huge opportunity that is not used in Sudan”*

Two of the participants felt that a greater emphasis on providing guidance around how to conduct community dialogues would help to generate demand for services and encourage voluntary contributions from those in the informal sector.

> *“But I think the biggest obstacle is. He’s a cultural one. And I think it. I think it if there’s a sort of groundswell, I guess of support I felt. Community and community engagement which would support that kind of universal health coverage. Fair access for everybody we all look after each other”*
>
> *“Why [are voluntary NHIF contributions so low]? Because there is no community dialogue guidelines. There are no guidelines for community engagement framework”*.
>
> *“I mean that’s the one thing we you know when you’re developing a central health benefits package; you know the first thing the guidance says … community engagement must start, you know. You engage your population, get to community. And that same goes for health financing…It was the one thing we couldn’t do because of the political sensitivities. And yet there are community groups already established. You know, many of whom were active in the the democratization of Sudan. Or the attempted democratization of Sudan in the in the 2019 initiative. Many of those groups exist and could be used as platforms for having the discussions that need to be had”*.

### 4. Purchasing and Service Delivery

#### 4.1 NHIF as purchaser and provider

While coverage and entitlement to service under the NHIF expanded with the inclusion of ‘poor families’, this did not necessarily translate to increased and improved access to service. There is confusion about which health services are covered by NHIF, and which services are purchased directly by the FMOH. In addition to acting as a purchaser of service from the MOH, the NHIF also has its own service providers without a clear demarcation between the purchase-provider functions. The FMOH adopted a policy to separate and refocus the roles of the NHIF as the purchaser of health services, and MOH as a provider of government health services, however, the implementation of this policy is incomplete (33, 34). As such, there are two state-backed, poorly integrated systems operating in Sudan.

> *“And the National Insurance Fund had sort of launched… When we looked from afar, it looked like it was almost a separate health system because the there was not a great connection between the National Health Insurance Fund- and state-funded health systems. There was a commitment to separating purchasing and providing and that the National Health Insurance Fund was essentially going to become the purchaser. And the ministry’s hospitals and facilities would be governed as a provider. But that policy hadn’t really been implemented”*.
>
> *“In Sudan here there are two modality of service provision under the context of NHIF. One of them is the purchasing from the facilities that affiliated to the Ministry of Health and the other is a NHIF facilities which it’s owned by the NHIF…. So now there is the problem of accessibility to services. Because of these facilities are not capable … there is a problem of integration of services”*.

#### 4.2 Inadequate service delivery

Budget capture by secondary and tertiary care seems to have made it difficult to purchase adequate primary care services in many areas. Desire to extend health coverage to these underserved populations may drive some of the instances where the NHIF strays from purchasing into direct service provision.

> *“There is a policy, there is that regulation, this commitment during the budget development that we are going to allocate so and so [to primary care], and we have to follow Abuja declaration on other. But when we come to the reality, we realise that most of the resources, most of the resources goes directly to hospitals and goes directly to the tertiary level which provide do you know services for only 20% of the population”*.
>
> *“ the health national health insurance fund is in some states they take a lot of a burden of giving the, even, they even, what do they call it? They establish the service. They build the the facilities which is not their role. It is our [MoH] role. Their role is to provide the service, you know, to pay for it, not to provide it… While we try to provide the service and prepare the facilities and train the people and do all that”*.

### 5. External Donor funding

#### 5.1 Verticality and fragmentation

Currently, 6% of health expenditure is from external donors, however it remains vertical and fragmented with different donor organization having different mandates and scope of work. Per participants’ accounts, budgets are developed in accordance with donor’s budgeting principles and priorities that do not always align with actual needs, particularly those of individual States, and do not reinforce integration efforts within the wider health system. Most donor funds are earmarked for specific programmes e, g. HIV, malaria, etc. which can create very fragmented funding streams with little integration into the overall system. Funds are mainly made to the FMOH despite the fact that most implementation occurs (or ought to occur) at State level. There is no accountability at State level to ensure funds are absorbed and activities implemented. However, weak State-level capacity has rendered donors hesitant to pay States directly but this has thereby resulted in a much-fragmented system of channelling funds to state level.

> *“we talked many times about the verticality we talked many times and all the time, I think now is the time to take action. We can’t go very well in immunisation but very bad in nutrition, very well in this, it is very bad. This will impact the overall output our total impact, the impact indicator of the health system So it’s just looked like fragmenting. This is not our effort. We provide more than that. So this verticality not affect the finance, only it affect the service delivery affect the health information and we have to use the hat of the health system thinking and not to focus only on the. So this is the very important the number three to address this verticality and time by time it’s a very good thing”*.

#### 5.2 Coordination of external donor funding

In the recent conflict, there is heavy reliance on donor funding from development partners. This has effectively diminished the government’s decision-making power and while there is some degree of coordination of how the funds will be spent, the locus of decision-making has shifted beyond the borders of Sudan. Participants asserted that coordination efforts between partners is also managed by an external organization, which creates a missed opportunity to include state and district level stakeholders and erodes the government’s ownership and oversight over health spending. There is a desire for joint planning and implementation between the government and donors, married with an acceptance that this is unlikely to happen. The fact that the government does not have control of all areas of the country compounds the issue.

> *“Joint planning, joint implementation, and joint reporting as well - I don’t think this is possible. For instance, to have joint budgeting, because yeah, it’s like, neither the government nor the partners would like to bring and pull their resources in one place, but it’s still. Having joint planning, joint implementation, for example, this might together reduce any inefficiencies might come up during the implementation. That’s one of the key issues”*.
>
> *“So, for me, for me in general, during humanitarian settings the control of government will come down and then because most of the time, sometimes the government cannot access areas of, conflict for instance. Access areas where RSF there and RSF cannot access the area of where SAF is controlling. So the control of government during humanitarian setting is usually less And that’s why what they are doing right now just to have this coordination mechanisms to make sure that things in the ground smoothly. For instance, UN agencies right now have huge shipments. From Khartoum from Port Sudan, through Aljazira to Kosti to be, send it to some of Kordofan states and also East Darfur But OCHA is leading the negotiation with SAF and RSF How to have. Safe movement of this logistics. Because there is no one government controlling on the ground. So, and that’s why the main, yeah, so. I think that’s, my sense. The government is just like coordinating. They don’t have the power to control everything and so on”*.

## Discussion

Perhaps surprisingly, our key informants felt that the greatest challenges to Sudan’s health financing came not from the recent conflict, but from underlying structural issues: the governance structure, revenue raising model, high self-financing payments for health, and fragmentation of funding sources. The main source of NHIF funds is from compulsory contributions from civil servants which constitutes only 13% of the population; the remaining funding of NHIF coming from the Ministry of Finance providing coverage for ambiguously defined ‘poor’ populations. The remaining proportion of the population are categorized under the informal sector who find no incentive to enrol under the NHIF. The considerable disparity between the States in terms of funding and capacity to raise revenue has led to stark disparities in health financing and funding of services. Lastly, external donor funds, while being crucial to the function of many key FMOH services, have fragmented the flow of funds to the health system and exacerbated inequalities between states. The total amount of money flowing through the system has fallen dramatically as funds have been diverted to defence spending, and external support has been withdrawn. However, service provision was already poor before 2023, and the vast majority of health spending was already out-of-pocket. As Sudan looks to the future, the great priorities are around developing community advocacy for risk pooling and essential health services, with much greater coordination and clarity around the roles of Sudan’s various federal and State purchasers and providers.

**Military coups** are not uncommon in low- and middle-income countries particularly in the African continent. As of 2022, 45 of the Africa’s 54 countries have suffered one or more coup attempts. On average, African states have suffered four coup attempts since their independence, with Sudan experiencing the most (a total of 17), including two in the previous year (35). This shows an increasing trend with 13 percent of African countries suffering coups in the prior two years, comprising a six-fold increase from just four years earlier (35). The long-standing political turbulence in Sudan has certainly taken its toll on the health sector, but research examining the effects of politics is scarce. Storeng and Mishra observed that studies on health systems strengthening while emphasising the technical and managerial of health systems scarcely gave attention to ‘the politics and social relations that shape health systems’ (36). While the WHO Commission on the Social Determinants of Health recognises that health inequities arise from ‘a toxic combination of poor social policies and programmes, unfair economic arrangements, and bad politics’ (37) there is a lack of recognition of public health emergencies that are of a more political nature as an existential threat to health systems. The case of Sudan is an exemplary representation of such circumstances.

**Health purchasing** can generally be defined as the allocation of pooled funds to the providers of health services(38). Ideally, a ‘strategic’ form of purchasing promotes efficiency and quality provision of services would be based on target population, national health priorities, and cost-effectiveness of selected interventions (39). The current service provision structure in Sudan is broad in scope and entitles its beneficiaries to a range of free healthcare, mainly primary care including medical consultations from PHC providers, general practitioners and specialists, laboratory investigations, and medical imaging (e.g., CT Scan and MRI). For medicines, users are expected to pay 25% of the cost. Despite this broad nature of coverage of the current package of health services delivered in Sudan, there are many challenges to its efficiency mainly related to funding which has to a large extent led to the high proportion of OOP expenditure in Sudan (33). A provider payment mechanism (PPM) i.e. the money which is transferred from a payer/purchaser to a provider of service as compensation for the delivery of the health services, must be agreed upon and institutionalised to ensure fair and equitable access to health services (40) while mitigating providers’ incentive to select healthier patients or over-providing of care (41).

In the case of Sudan, there was no clear distinction between purchasing and provider functions of the NHIF. In 2019-2020, and commissioned by the WHO, a Health Benefits Package (EHBP) and a Provider Payment Mechanism (PPM) for the Health System of Sudan was designed to support the overall aim of health financing reform in the country following the establishment of its new transitional government. The project proposed that the NHIF’s role in the financing system should be confined to ‘payer’ or purchaser, while service delivery relegated to the Ministries of Health; a proposal much welcome by most of the study participants. Under the proposed separation of payer and provider functions, each SMOH would be considered a single provider of MOH services within each State, with a separate agreement with NHIF for the provision of health care for the state population. Furthermore, the project proposed establishing a standard and consistent PPM model used by NHIF to pay SMOH for service delivery while preserving state flexibility through mixed payment models for SMOHs to use to fund different SMOH health care outlets. The proposal also noted the various challenges this would inevitable face including the inequity in epidemiological and economic status across states (33); in addition to the political and security challenges faced by conflict-prone states that would hinder effective service provision (33, 34). The political instabilities, coup and most recently the war in Sudan has hindered implementation.

**Risk pooling** or lack-there-of, was a prominent theme in the results of this study. The current structure of the NHIF caters to primarily those in the formal sector who constitute only 13% of the population. Services that are currently delivered under the NHIF are not consistent or comprehensive and while they may be free in theory, many services may not be available at all, or be of poor quality (33). The unattractive structure of service delivery has proved ineffective in attracting subscribers from the informal sector to the NHIF. Furthermore, providers charge co-payments or full payments for access to services to supplement the funds they receive from the SMOH, NHIF, and NGOs

(33)which places a high burden for OOP costs and limits the effectiveness of risk pooling intended by the establishment of the NHIF (42). In fact, a recent study showed that insured and non-insured citizens had similar OOP payments in public healthcare facilities that led to catastrophic expenses which not only affected the poor but also the highest quintile of the population (42). The same study suggested that the NHIF has ‘failed to protect people against health-related financial hardship’ and indicated many factors that have resulted in this failure including low coverage of the informal sector, low quality of provided health care services provided to insured people, and most crucially, lack of coordination between the NHIF and FMOH to establish efficient and effective financial agreements to protect people from catastrophic health expenditures (42, 43).

Risk pooling refers to the subsidization the health population extends to the ill and spreading the financial risk of costs borne from seeking healthcare among the population (45, 46) Lack of risk pooling typically results in a negative correlation between health and costs of care; costs of which, in the absence of risk pooling, would be endured largely by the ‘sicker’ population. (45, 47). This is inconsistent with the objective of financial protection and equity of access to services in relation to need (45). Pools can generally be categorized based on mode of participation into on compulsory, automatic or voluntary. Typically, pools comprised of compulsory coverage is compulsory or automatic for all population groups and have a more diverse mix of health risks. Conversely, pools comprised of voluntary membership, individuals with higher risks are more likely to enrol than people with lower health risks leading to an inadequate diversity of healthier and sicker people, and the costs of care would be significantly higher than for the average in the same population limiting potential and efficacy of risk pooling (45, 48). In the case of Sudan, the lack of incentives to enrol in the NHIF, with OOPs remaining similar for those enrolled and those not enrolled, results in lower levels of risk pooling, and burden of health expenses borne by the less advantaged populations.

However, the extent to which a health system effectively achieves its risk pooling objective is largely conditional on the amount of revenues raised among other factors (45, 49). The capacity of a country to raise resources depends on its economic status and national income and can be classified as either compulsory of voluntary; the former relying primarily on taxes and other governmentally levied charge; and the latter mostly sourced from OOPs (50). The Federal Ministry of Finance is the major contributor in financing NHIF (72%-mainly from tax revenues), followed by the parastatal organizations (12.7%), while households’ contribution is at approximately 9% (51). A tax-based or tax-funded system, such as the NHIF, is very sensitive to economic and fiscal policies and shocks to the national economy and ‘fiscal space for health’ i.e., how much a country can spend on health depends on how much revenue it can raise from tax, how much is available to spend by government and finally what share of this can be spent on health (50); rendering sustainability questionable at best.

At present the main source of health financing is the NHIF compulsory premiums which represent only 13% of the population while the remaining sources are the Ministry of Finances’ contribution towards the ‘poor’ populations and OOPs. The study participants almost unanimously expressed their strong support for a mixed sourcing of revenue raising i.e., increasing the NHIF pool by increasing the enrolment into the Fund while ensuring taxes are used to ‘top-up’ the Fund resources. This is line with many studies that collectively agree on ‘a serious need for additional financing sources’ to the NHIF (52). The mixed source of financing for the NHIF would limit- to some extent-the reliance of the NHIF on tax subsidies and thereby the implications of financial and economical blows to the Sudanese economy which-as witnessed in the most recent conflicts-severely depress the government’s ability to contribute to the NHIF (52).

A mixed or blended form of revenue raising requires significant increase in citizen contribution into the NHIF. As highlighted by the study participants, there is a lack of trust between communities and the NHIF with respect to efficiency and quality of services provided. Community engagement is a powerful and necessary tool in expanding NHIF subscription among the mostly informal sector population. Community engagement not only serves the purpose of establishing accountability on NHIF and MOH, but also relegates decision-making processes to the local level (53). Community engagement has not been a priority for the FMOH and no guidelines for community engagement were developed to encourage dialogue. It was only recently a community engagement dialogue project was completed in North Darfur which was the first to develop a ‘Community engagement Dialogue Framework’ and it remains to be context-specific. Strategies suggested to improve citizen participation in NHIF have included active recruitment, simplifying the process of enrolment, reaching citizens in their places of work and even making health insurance a condition for attending public healthcare facilities (51) although in light of the limited service availability, this would be very counterproductive and would rather increase inaccessibility.

The stark variation between the different States in terms of health system capacity and ability to raise funds is consequential when designing reform policy and developing a blended or mixed PPM. The federal and state designation of roles and responsibilities albeit as opaque as it is-also creates complication – alongside the political complexities and ethnic sensitivities that exist between states. There is a pivotal need to develop allocation mechanisms that account for population size, population needs, state capacity to raise tax, and state health system capacity to provide services - in addition to accommodating for special needs, particularly for conflict-prone states such as the Darfur states. This should ideally be supplemented by funding from NHIF at federal level; this approach would be context-sensitive and specific, observant of state level needs while allowing for state flexibility in raising revenue and self-identifying needs; thus, weaker and ‘poorer’ states would be more eligible for federal support. The federal level is instrumental in aligning these allocation mechanisms while ensuring political feasibility of application.

A third source of resource-critical for many LMICs health systems are external donor funds. While external financing is critical for many LMICs, external financing has come under scrutiny in recent years for infringement on country ownership, fragmentation of service delivery, inefficiency and diverting experienced civil servants from government positions (54, 55). A recent review of health financing in fragile and conflict-affected setting found that external aid coordination was the largest single topic within the themes of the study, likely reflecting the strong influence of external players (56). Similarly, Sudan is heavily reliant on external donor funds for many key health services e.g., HIV, TB and malaria through the Global Fund; vaccinations through Gavi the Vaccine Alliance; and broader health service support through the World Bank among many others. Donor funding flows directly to the FMOH which then funnels the grants/funds to states and localities. Partner-funded programmes are not well integrated with local or national services; for example, an ANC health facility may be extremely underfunded but may receive excellent support in its vaccination programme, creating a confusing picture when it comes to overall health outcomes. This verticality does not serve reform purposes and rather creates further division and opacity in terms of funding streams.

The verticality of donor-funded programmes is well documented in the literature, and despite the agreed need for better coordination between donors and recipients, the literature tends to focus on efficiency and effectiveness rather than duplication and ownership.(57). In the recent crisis, and with the almost complete collapse of FMOH functioning, external funding has been essential to maintain many basic services in Sudan. In the absence of strong FMOH leadership, this has meant that donors have been left to operate in complete independence and will full autonomy, with little to no influence from the FMOH. As the conflict has evolved, the FMOH has regrouped, reorganized and began to operate from remote locations, however, there is a great need for improved coordination between FMOH and donor organizations with national bodies taking on more stewardship roles. Study participants reported an abundance of mechanisms designed to improve coordination, however effective implementation is where they have all come short. The political instability in the country in recent years has further fuelled this lack of progress with considerable reliance on communities and grass-roots movements such as resistance committees.

## Conclusions

The study has highlighted the deep-rooted challenges in the health financing system in Sudan. Despite the conflict, Sudan retains the basic health financing apparatus in the form of state and federal ministries, a basic national insurance fund, and basic health services. These elements can be reformed and leveraged to improve resilience of health financing in the face of the current and future crises. Whilst the current NHIF is clearly flawed, it represents an important vehicle for extending financial risk protection and the extension of essential health services. It is currently primarily funded through government taxes and contributions of premiums from the formal sector. The most pressing need is to include informal sector workers. OOP remains among the highest in the region, despite the country being one of the world’s least developed countries. Better pooling policies and revenue raising mechanisms are needed to improve equitable access to healthcare in Sudan. The vast discrepancy between states in terms of state capacity and health system capacity demands a tailored approach to allocating mechanisms between federal and state level funding. Lastly, external donor funded programmes must be better integrated into PHCs and reduce fragmentation within the broader health system. Based on our findings, we propose the following policy options to develop a more resilient health financing structure in Sudan:

## Recommendations

### Reform of existing NHIF fund

Despite the flaws in its design, the NHIF has shown some degree of stability in the face of major crisis. Efforts can be made to reform the NHIF to improve equity and efficiency and support the resiliency of the Sudanese health financing model. The reform policy must include:

- **Develop a blended system of NHIF revenue raising** that combines tax and premium based insurance contributions. This has multiple benefits: a) increases solvency and allows for less reliance on tax contributions lessening the impact of fiscal austerity and economic decline; b) improved risk pooling and reduces the moral hazard and adverse selection effects; c) more equitable form of coverage.
- **Develop a community-targeted strategy to encourage enrolment of those working in informal sectors.** The FMOH/NHIF should leverage momentum from existing grass root and community movements that have been central and instrumental in the covid-19 response and during the ongoing conflict in providing health services and awareness raising in their community engagement strategies.
- **Establish clear definitions of what constitutes informal sector and ‘poor’ populations**: Despite the NHIF’s annual update of selection criteria, for example, by adopting the World Bank (58) international poverty line of $2.15 per person per day using 2017 PPPs. Since 2014, there are no official poverty statistics (58) and data collection methods are not robust enough to collect needed information to categorize and determine eligibility criteria. A robust and preferably digitalised data collection method would support not only identifying eligible participants, but broadly speaking, would support more efficient tax raising efforts, particularly at state level. FMOH and NHIF should leverage existing partner-funded (e.g., UNICEF,GAVI and the Global Fund) DHIS2 strengthening programmes (5, 59) to inform NHIF eligibility criteria.
- **Innovative pooling of funds within the NHIF for emergencies and crises**: An innovative pooling mechanism that includes donor funds, private sector and other religious institutions could support the existing emergency reserve within the NHIF. This must include clear and specific trigger mechanisms under the control of FMOH and NHIF, and allocation specific instructions. For example, the World Health Organization in Sudan is working closely with the FMOH to develop an emergency prioritisation strategy to identify essential interventions in cases of emergencies. There must be strong alignment with these two siloed activities to avoid duplication of efforts and maximize efficiency of any reserved or emergency funds.

### Strengthening Policy Implementation

The FMOH in collaboration with development partners has a multitude of existing policies, plans and strategies for financial reform. However, these have not been coupled with implementation and monitoring plans. Moreover, there is no framework to hold underperforming partners accountable. To improve implementation and accountability during the current crisis and beyond, the following is suggested:

- **Invest in capacity building particularly at state level**: Most states do not have the capacities to inform their health financing policies and/or implement them. Led by the FMOH, institutionalizing a structure similar to the technical steering group at state levels could support local training and capacity building efforts to accelerate implementation.
- **Improved coordination with development partners to create synergies with existing programmes to better respond to emergencies**: in a country such as Sudan which is prone to armed conflict and natural disasters, more effort must be placed in anticipatory coordination with external donors and partners to align priorities and resources. At the moment there are emergency ad-hoc/monthly/bi-weekly ‘National Health Sector Partners’ Forum/Cluster’ meetings between FMOH/SMOH, primarily led by development and humanitarian partners working from various locations inside and outside of the country. This is a good strategy to coordinate collective efforts and prevent duplication, that can be leveraged post-conflict by instituting biweekly/monthly cross-sectoral health management committees but this must be led by the Planning Directorate at FMOH- and include SMOH, development partners, and civil society- to ensure ownership and integrated, streamlined processes(60).These committees will regularly review national policies and oversee implementation progress.
- **Strong governance for health**. For the above recommendations to be successful, strong leadership and commitment is necessary from the Ministers at head of FMOH and Federal Ministry of Finance. Policymakers within the Planning Directorate at the FMOH should start by making the business and economic case for government investment in health to advocate for more spending on health (33) to be discussed and disseminated with relevant stakeholders.

## Data availability

No data beyond those presented in the manuscript are available. This is to protect the interviewees.

## ANNEX 1

Interview Guide

**Q1: As you reflect on recent events, can you tell me more about the current health financing landscape? how financing policies are made and how the multitiered structure of the health system works?**

Probing questions:

- How are those decisions made at federal level?
- How do SMOH access funds? How is the proportion/share of each state determined?

**Q2: In the event of a major political crisis, we see the health sector be among the first to be severely affected, are there mechanisms in place to mitigate this?**

- Is this mechanism a standard policy/protocol employed by the FMOH? How are these decisions made?
- How does the aftermath of such events affect this decision-making process?
- How do you assess the effectiveness of such measure? How successful have they been in mitigating these effects?

**Q3: What has happened to health financing in Sudan since April 2023?**

- How has this impacted service delivery?
- How has this impacted equity?
- How has this impacted financial protection?
- How has this impacted quality?

**Q4: who are the major actors in the health financing space at the moment?**

- What are their interests
- What decision-making power/influence do they have?

**Q5: what could/should be done now to release needed funds?**

**Q6: in the longer term how could we safeguard funds?**

- How feasible is this option?
- Who are the relevant actors, and how does this align/misalign with their interests?
- What would need to be in place for it to be a feasible and sustainable solution to mitigate the impact of negative political events/crises?

## Annex 2: COREQ checklist

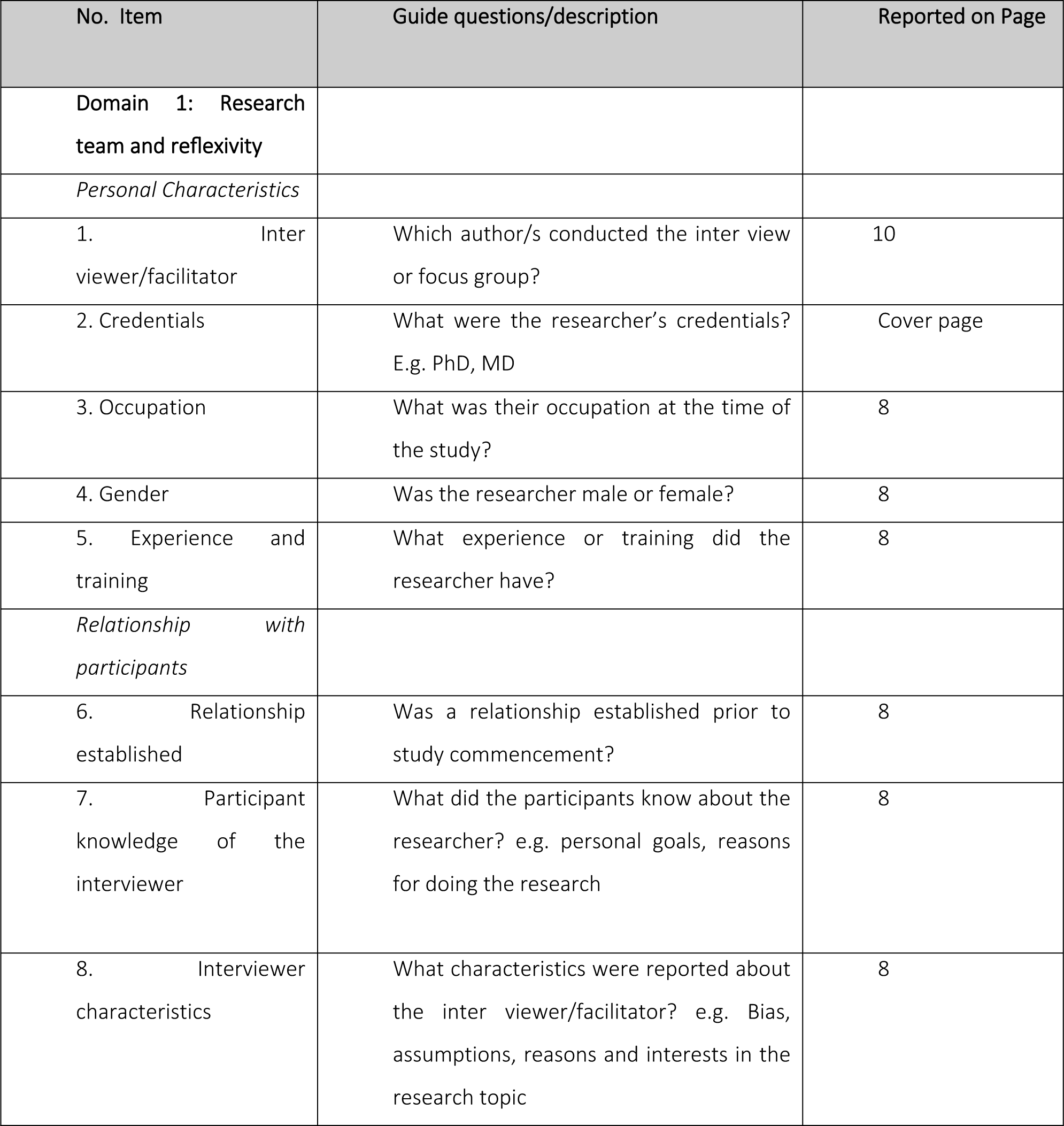

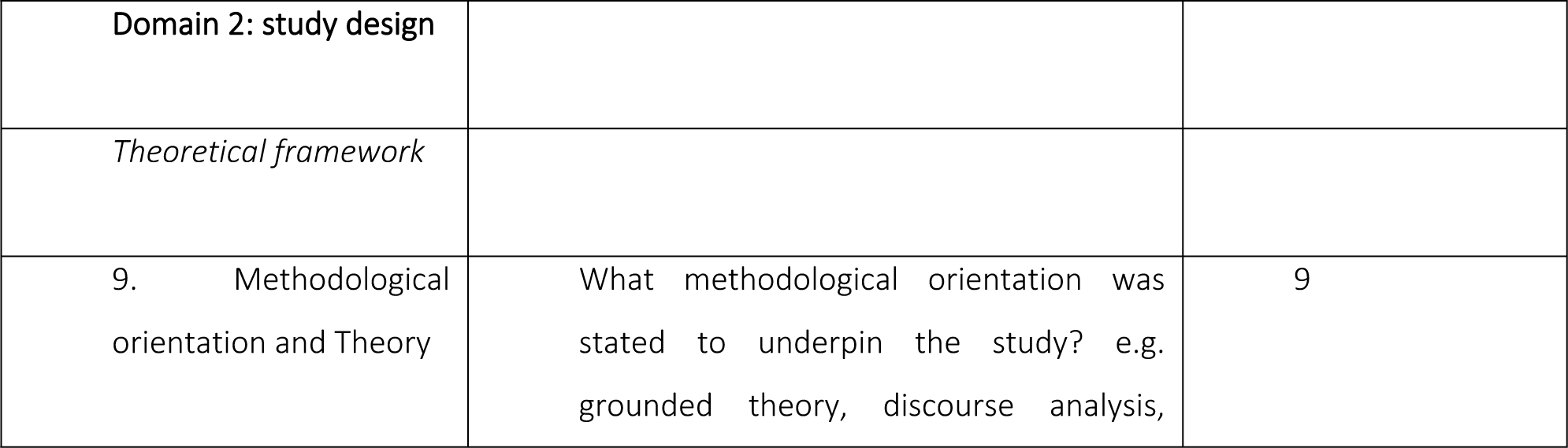
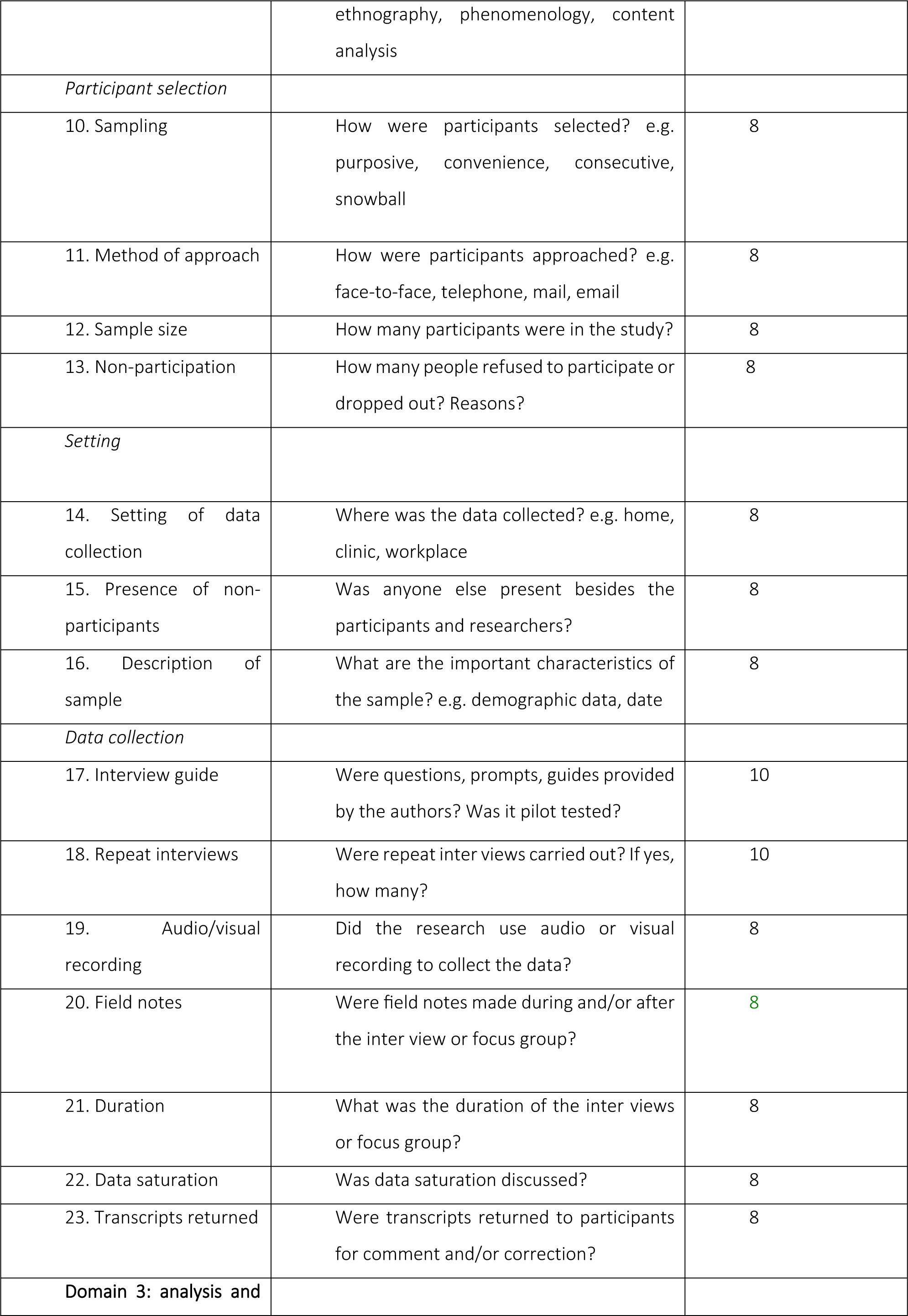
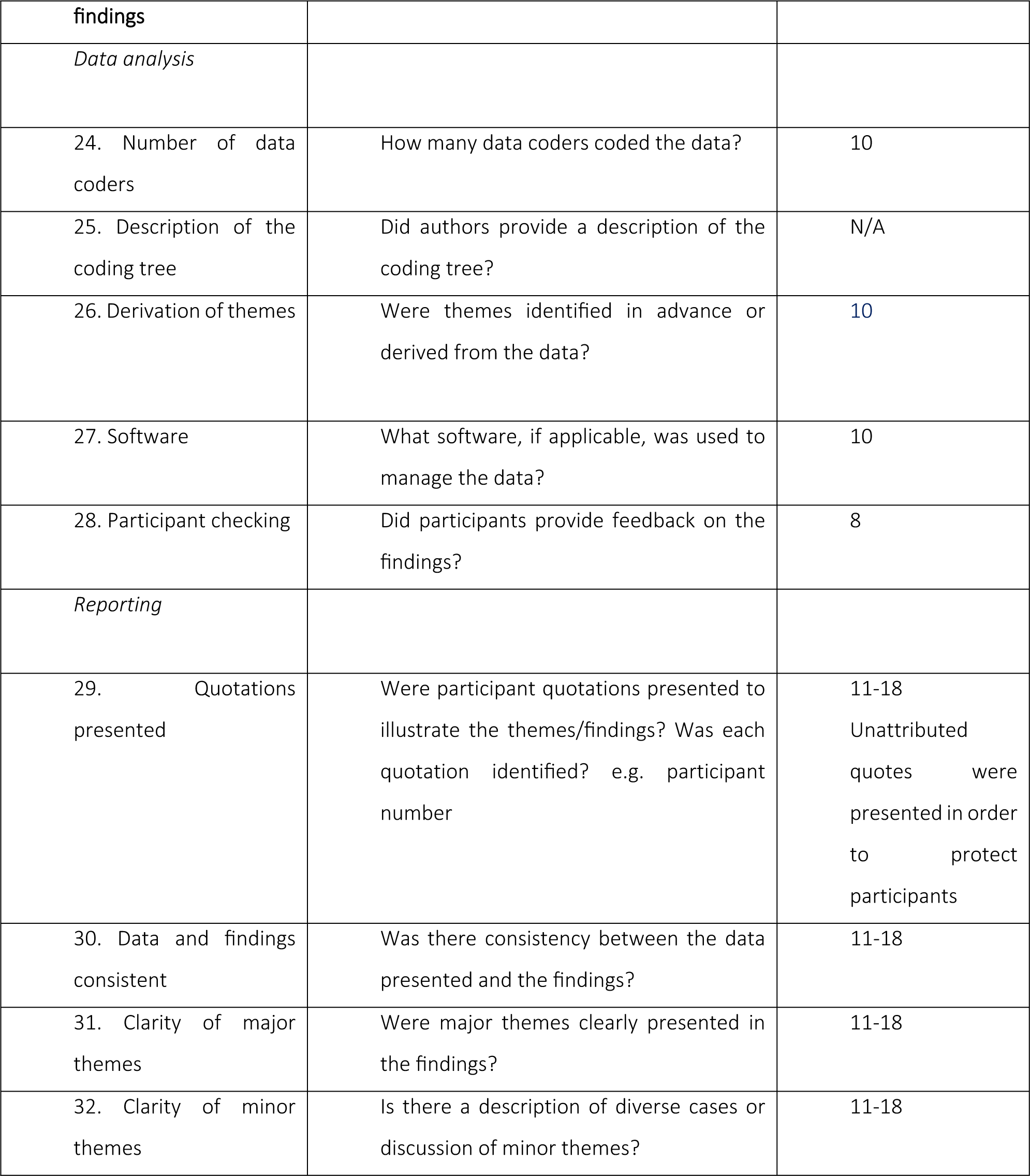

